# The Effect of Combined Respiratory Muscle Training (cRMT) on dysarthric speech following single CVA: A retrospective pilot study

**DOI:** 10.1101/2021.05.01.21256463

**Authors:** Robert J. Arnold, Nina Bausek, Christopher S. Gaskill, Tarek Midani

## Abstract

**Background:** Dysarthria frequently occurs as a result of stroke and adversely impacts speech sound production, making it more difficult for the listener to understand what the person with dysarthria is attempting to communicate. This in turn may lead to social isolation, depression, and increased cost of care. Some studies have underscored the importance of respiratory muscle strengthening as it relates to improvement of speech intelligibility, However, the benefits of respiratory muscle training on dysarthric speech following stroke have not been explored. This retrospective investigation examined the effects of a combined Respiratory Muscle Training (cRMT) protocol upon speech intelligibility in persons post single cerebrovascular accident (CVA).

**Methods:** The clinical data of 10 patients who requested speech therapy at a pro bono outpatient clinic for the diagnosis and treatment of dysarthria following a single stroke was utilized for this study. The intervention group was treated with three 5-minute sessions with cRMT each day for 28 consecutive days. The control group received no cRMT and no other therapeutic exercise intervention during the time period. Respiratory function and speech intelligibility were assessed pre- and post-intervention in terms of peak expiratory flow, subject self-perception of intelligibility, and word level intelligibility.

**Results:** After 28 days of cRMT, the intervention group exhibited significant gains compared to the control group in peak expiratory flow (PEF) (IG: 73.12% vs CG: 4.66%), Self-Perception of Intelligibility (IG: 72.38% vs CG: 0.83%), and the word task of the Assessment of Intelligibility of Dysarthric Speech (AIDS) (IG: 43.92% vs. CG: 0%).

**Conclusion:** These retrospective data demonstrate cRMT is a feasible and effective treatment for improving breath support and speech intelligibility in persons with dysarthric speech after single stroke.

## Introduction

Approximately 20 to 40 percent of the 5 million stroke survivors per year in the United States of America have a motor speech disorder known as dysarthria (Arboix et al., 1990; Enderby & Emerson, 1996; Kunst et al., 2011; Lawrence et al., 2001; Lloyd-Jones et al., 2010). In the United Kingdom, about 1 in 53 persons (1.2 million) in the total population are stroke survivors, of whom 30 to 40% have had dysarthria at some stage (Miller & Bloch, 2017).

Dysarthria causes distortions of speech sounds, decreasing the ability of the listener to comprehend the content and emotion messages the speaker intends to convey. Severe dysarthria can even render the affected person’s speech totally unintelligible.

Stroke-induced dysarthrias may restrict social interactions, leading to isolation and depression. However, the severity of dysarthric speech is not always congruent with the psychosocial impact, as even mild dysarthria may have a significant impact (Brady et al., 2011). Dysarthria can result in impaired self-identity, dysfunctional relationships, emotional disruptions, and even perceptions of being stigmatized (Dickson et al., 2008). In contrast to the evidence for the psychosocial impact of dysarthria following stroke, the socioeconomic and financial burden appears largely uninvestigated. Although the cost of stroke and dysphagia is well documented, little investigation has been conducted with regards to the personal financial hardships and societal financial impacts of dysarthria (Wilson, 2012).

Lastly, personal clinical observations have shown that dysarthria can put persons at risk for untoward treatment outcomes when unable to communicate their signs & symptoms to a physician, surgeon, or other health care provider.

Dysarthria can be classified according to the Mayo Clinic taxonomy into flaccid, spastic, ataxic, and some mixed types of dysarthria (e.g. spastic-flaccid) in the stroke population. (Darley et al., 1969a, 1969b, 1975; Johns, 1985). Dysarthria subtype classification can also indicate the location of the stroke damage in the central nervous system (CNS). Flaccid dysarthria typically results from lower motor neuron damage, and spastic dysarthria from upper motor neuron damage. Identification of dysarthria subtype can also guide treatment selection.

The clinical characteristics of dysarthric speech following stroke include rate of speech, prosody, resonance, phonation, articulation, and overall intelligibility. These factors can be evaluated through perceptual, acoustic, manometric, and palatographic means. One commonly used clinical tool with adult populations is the Assessment of Intelligibility of Dysarthric Speech (AIDS) (Yorkston & Beukelman, 1981) which evaluates speech intelligibility at the word level and sentence level and provides a percentage of the intelligibility of speech at both levels. Objective measurements of lip, tongue, and jaw muscle strength and endurance, as they relate to speech and swallow functions, may provide additional insights into function and dysfunction of relevant muscle groups. Several recently developed technologies provide tools for specific evaluation (Arnold, n.d.; Arnold & Mann, 2004; Robbins, n.d.; Robin & Luschei, n.d.).

Early evidence presumed that the most limiting factor in breath support for speech in the presence of dysarthria was insufficient valving of the exhalatory airstream (Hardy, 1961, 1967). This is supported by the realization that the use of a relatively high lung volume at the onset of speech could improve the intelligibility of speech clarity (Hardy, 1968). Furthermore, some clinical researchers have asserted that if a given patient can continuously generate subglottic air pressure of 5 cm of H_2_O for at least 5 seconds or more, then the respiratory system is likely to be adequate for supporting speech efforts (DWORKIN & P, 1991). Use of this pressure generating capability alone may offer a limited perspective, as persons with neuromuscular respiratory deficits may be able to perform this simple static task, but still be unable to sufficiently perform the complex, dynamic respiratory and articulatory movements required for the demands of speech.

In recent years, data regarding normal and abnormal speech breathing has become available enhancing the current fund of knowledge pertaining to how impaired articulation may result not only from motor impairments of the muscle of the articulatory structures and muscles of the oral cavity, but also motor impairments of the muscles of respiration resulting in inadequate initiation and/or maintenance of subglottal pressure in connected speech at various levels of loudness (Hixon & Hoit, 2005; C. Sapienza & Hoffman, 2020b; Titze, 1992). Many neurologically impaired patients experience significant difficulty in the consistent regulation of their speech breathing (Bateman & Mason, 1984). Stroke has been identified as a common cause for disorders of respiration (Howard et al., 2001; Rochester & Mohsenin, 2002). Although physiological disorders of respiration resulting in respiratory muscle weakness (RMW) have been identified as being a contributor to loss of intelligible speech following CVA.[1], another factor which often further exacerbates the severity of respiratory muscle weakness after stroke is the effects of bedrest, which has been demonstrated to result in sarcopenia (disuse induced atrophy of skeletal muscles) at a rate of up to 40% within the first week of being bed bound with further loss of skeletal muscle strength at a rate of 12% per week (Jiricka, 2009; Topp et al., 2002). Prolonged bed rest alone has been correlated to increase the risk of developing respiratory tract infection after stroke as persons with stroke who are bed bound for 13 days or longer are 2 to 3 times more likely to have a respiratory tract infection when compared to healthy individuals who are still ambulating (Halar, 1994).

There is limited evidence regarding the effectiveness of treatment interventions of dysarthric speech (Sellars et al., 2005). Traditional approaches have included coaching of slow rate, breathing exercises, over-articulation, and active resistance oral range of motion tasks.

Newer approaches include Lee Silverman Voice Therapy (LSVT) (Mahler & Ramig, 2012; L. Ramig et al., 2018) and expiratory muscle strength training (EMST) (Jones et al., 2006), as well as some emerging concepts with augmentative and alternative communication (AAC) systems and voice banking including VOCALiD (Beukelman et al., 2007; Mills et al., 2014; *VocaliD – Your Voice AI Company, Bringing Things That Talk to Life*, n.d.; Yamagishi et al., 2012). Respiratory muscle weakness (RMW) has been found to be prevalent in patients with dysarthria and/or dysphagia following stroke, and amenable to respiratory muscle strengthening (Pollock et al., 2013). More recently, research has revealed the importance and effectiveness of respiratory strengthening for the treatment of dysphagia, dysphonia, and dysarthria (Arnold et al., 2021; Arnold & Bausek, 2020; Chaitow et al., 2014; C. Sapienza & Hoffman, 2020a, 2020b; C. M. Sapienza & Troche, 2011). A pilot study using respiratory muscle training (RMT), strengthening both the inspiratory and expiratory muscles, resulted in improvement of several aspects of RMW as it pertains to dysarthria and dysphagia following stroke, including fatigue, respiratory muscle strength, lung volume, and voice clarity (Liaw et al., 2020). One recent study showed expiratory muscle strength training (EMST) to have a positive impact upon expiratory muscle strength but did not statistically correlate with improved speech sound production in persons with multiple sclerosis (Chiara et al., 2007). A study examining the effects of expiratory muscle strength training in persons with Parkinson’s disease (Darling-White & Huber, 2017) resulted in preliminary evidence of EMST improving speech breathing. A recent systematic review of the literature regarding interventions used to improve pulmonary function after stroke found evidence of respiratory muscle training to be useful for improving both inspiratory muscle strength and expiratory muscle strength after stroke (Menezes et al., 2018).

Respiratory muscle weakness (RMW) is a significant contributor to dysarthric speech independent of the subcategory after stroke. Respiratory muscle training has been shown to have a direct effect on RMW and is considered a safe and useful therapy approach for improving RMW after stroke. As little to no information exists regarding the effects of combined inspiratory strength training and expiratory strength training in a single exercise upon speech intelligibility following stroke, this retrospective pilot study investigates the effects of combined Respiratory Muscle Training (cRMT) upon dysarthric speech following single stroke (Chiaramonte et al., 2020). In order to assess the effects of cRMT on speech clarity and breath support, this study investigates the effects of an exercise program using a combined resistive IMST and EMST device (THE BREATHER ®, PN Medical Inc., US Patent Number 4,739,987) (M. K. Nicholson, 1988), which allows for strengthening of the muscles of inhalation and exhalation in the same breath cycle (Shaikh et al., n.d.; Shaikh & Gunjal, 2019). This retrospective pilot investigation analyzes the effectiveness of a 4-week cRMT program, which has been shown to have positive effects on neurogenic dysphagia (Arnold & Bausek, 2020) and neurogenic dysphonia (Arnold et al., 2021), on the improvement of speech intelligibility and peak flow in stroke patients diagnosed with dysarthria following a single cerebrovascular accident (CVA).

## Methods

The source data for this retrospective study was collected at a *pro bono* clinical speech-language pathology (SLP) clinic, which was run by the Office of Hispanic Ministry at a Catholic church in the greater Birmingham area of Alabama between 1998 and 2010. This study was granted an Institutional Review Board (IRB) exemption by the Western Institutional Board. All participants included in the study presented with dysarthria following a neurologist confirmed single CVA. The average amount of time between the onset of each participant’s stroke and referral to the pro bono SLP clinic was 4 weeks. As there was a lag between diagnosis and an available treatment opportunity, patients were offered a choice to be instructed in cRMT in the interim, which included explanation, demonstration, and return demonstration. Patients who accepted the offer underwent a four-week home-based intervention of cRMT using The Breather. Patients who chose to participate in the cRMT program were included in the intervention group (IG), while those who chose not to use the device while waiting for treatment were considered as the control group (CG) for analysis.

Participants included in the analysis had no prior neurological history, no history of dysarthria prior to their CVA, no pharmacological or surgical interventions, and no additional therapy for speech, swallow, or voice during the study period. Furthermore, no other allied health therapy services which may have resulted in some degree of improvement of articulatory, vocal fold, or respiratory functioning (e.g. occupational therapy, physical therapy, respiratory therapy, music therapy, etc.) were administered during the study time period. Lastly, there were no changes in medications noted (e.g. changes in dosage, discontinuation, addition, etc.) during the study time period. Persons documented to have had an extension of CVA were excluded. Any patients who failed to meet one or more of the inclusion criteria were excluded from this study. The participants’ demographic data were reported by age and sex. Additionally, the specification of the hemisphere of CVA was reported.

### cRMT Intervention

The intervention resulting in the data used for this retrospective study focused on combined respiratory muscle training (cRMT) and was administered using THE BREATHER ®. Patients were instructed to sit upright, and to forcefully inhale and exhale through the device using diaphragmatic-abdominal breathing. If necessary, patients were advised to use a nose clip. If patients were unable to maintain a tight lip seal using the mouthpiece of the cRMT device, they were presented with the option to utilize a CPR (cardiopulmonary resuscitation) facemask and received appropriate training. The duration of intervention was 28 days (4 weeks), and included three sessions per day. One session per week was supervised by a volunteer speech-language pathologist who was present for each of these weekly sessions. Each treatment day consisted of 3 sessions per day of up to 5 minutes of cRMT per session, as tolerated, judged by the patient’s report of fatigue or dizziness and the clinician’s assessment of patient fatigue. cRMT intensity was defined as the highest tolerated settings for both inhalation and exhalation. Settings for inhalation and exhalation were set independently from each other. Patient compliance and adherence to cRMT were assessed via patient communication during the weekly supervised intervention sessions. During the study duration, the control group patients by their own choice received no treatment.

### Assessments

To assess pulmonary function, we assessed expiratory peak flow (PEF) using a peak flow meter (Spir-O-Flow Peak Flow Pocket Monitor, Spirometrics) by taking the average of 3 trials as there is a correlation between expiratory peak flow and the pulmonary function required for speech intelligibility. Participants’ self-perception of their speech intelligibility was assessed using a visual analogue scale (VAS) with a 100 mm horizontal line where “0” on the far left represented “My family and friends don’t understand anything I say” and “100” on the far right represented “My family and friends understand everything I say”. Participants judged their own speech intelligibility both pre- and post-treatment period by placing a vertical hash mark somewhere along the horizontal line. The clinician then measured the distance in millimeters from the left end of the horizontal line to the hash mark, so the speech intelligibility self-perception data is presented as a number from 0 to 100. Speech intelligibility at the word level was determined by administering the Assessment of Intelligibility of Dysarthric Speech (AIDS), which is an instrument designed to quantify speech intelligibility at the single word level and the sentence level (Yorkston & Beukelman, 1981). For the purposes of this retrospective pilot study, only the single word intelligibility was assessed for each participant. The tasks to assess speech intelligibility at the single word level utilizing the AIDS allows the examiner to require the dysarthric speaker to either read single words aloud or, whenever this is not possible due to comorbid visual-perceptual deficits and/or inadequate premorbid reading abilities, to imitate the examiner’s spoken model of each word assessed. Of these two methods, the former is considered to be the preferred method. Whichever method was employed in the initial assessment for a given participant on day 1 was also used for follow-up assessment on day 28 for that same participant. Regardless of method, each participant’s spoken samples were audio-recorded onto a digital recording device in a quiet room. The recorded samples were then provided to 2 blinded judges who were speech-language pathologists, neither of whom were the examiner. Each judge was provided with a data worksheet copied from the AIDS test manual. The judges were required to listen to each of the 50 words recorded by a given dysarthric speaker. As they listened to each word, the judges would select one word from a field of 12 written possible words on the data worksheet. Once completed, the examiner, using a key of what the actual word was for each of the 50 samples, would calculate the percentage of words correctly understood, which represented the speech intelligibility of the given dysarthric speaker at the word level at that one point in time. Next, the scores of the two judges were averaged to derive the average intelligibility of speech at the word level for data analysis in this study.

### Data Analysis

Group level statistical analysis was utilized in spite of the limited N to ascertain the presence versus absence of a pattern suggestive of a positive therapeutic effect of the cRMT intervention upon the expiratory peak flow, speech intelligibility, and patient self-perception of improvement. The primary research question was examined with one multivariate 2 × 2 factorial analyses of variance with repeated measures. The between-subjects independent variable was the experimental group (control vs. intervention) and the within-subjects independent variable was time (pretest vs. post test). The three dependent variables were the average of the three peak flow readings, the self-perception of intelligibility, and the word-level intelligibility task.

Univariate analyses followed significant multivariate analyses. The primary focus of the analysis was to determine if there was a significant difference between the control condition and the experimental condition over time due to the intervention. The multivariate interaction effect of time by the experimental group determined whether the two experimental groups changed from the pretest to the post test.

## Results

### Demographic and Clinical Data

This study includes ten participants. Demographic data including gender and age are outlined in table 1. The median age was 69 for the control group and 71 for the intervention group, with no significant difference between the groups (pValue: 0.85).

**Table I:** Distribution of gender and classification of dysarthria.

|  | **CG** | **IG** | **Total** |
| --- | --- | --- | --- |
| **n** | 5 | 5 | 10 |
| **Gender** |  |  |  |
| Female | 3 | 4 | 7 |
| Male | 2 | 1 | 3 |
| Median Age | 69 | 71 |  |
| **Classification** |  |  |  |
| Spastic Dysarthria | 2 | 3 | 5 |
| Flaccid Dysarthria | 2 | 1 | 3 |
| Ataxic Dysarthria | 1 | 1 | 2 |

**Legend:** This table shows the gender distribution and median age of the intervention group (IG) and control group (CG), as well as the distribution among the dysarthria subtypes.

Table 1 also describes the classification of dysarthria identified in the subjects. According to diagnosis of the referring neurologist, of the five participants in the control group, one patient had a left hemisphere upper motor neuron (UMN) infarct which correlated with the presence of spastic dysarthria; one patient was diagnosed with a right hemisphere UMN insult and spastic dysarthria; two were diagnosed with lower motor neuron (LMN) infarctions in the brainstem and flaccid dysarthria; and one was diagnosed with a cerebellar infarct and ataxic dysarthria. Out of the five participants in the intervention group, two were diagnosed with left hemisphere UMN infarctions and spastic dysarthria; one was diagnosed with a right hemisphere UMN insult and spastic dysarthria; one was diagnosed with LMN infarct in the brainstem and flaccid dysarthria; and one was diagnosed with a cerebellar infarct and ataxic dysarthria (see Table 1). None of the participants in this study presented with hypokinetic, hyperkinetic, or mixed dysarthria. All participants reported English as their primary language.

### Main Results

#### Pre-test analysis

The two experimental groups were compared on the pre-test measures (Average peak flow reading, VASA, and Word Task) using multivariate analysis of variance (MANOVA). The multivariate test was not significant, (p = .853). The two groups were not significantly different from each other on any of the pre-test variables.

#### Main analysis

Table 2 outlines the findings from the main assessments of peak expiratory flow, speech intelligibility and word-level intelligibility. We first examined whether there was a main effect for time for the set of three dependent variables and second whether there was a difference between the control and intervention groups over time (Complete results from the statistical analysis are available in supplementary material).

**Table II:** Pre cRMT and post cRMT assessment of average peak flow, self-perception of intelligibility, and word-level intelligibility

|  |  | Pre cRMT |  | Post cRMT |  |  |  |
| --- | --- | --- | --- | --- | --- | --- | --- |
|  | Experimental Group | Mean | SE | Mean | SE | Percent Change | p in between groups |
| Average Peak Flow (L/min) | CG | 85.80 | 8.33 | 89.80 | 8.16 | 4.66% | .001 |
|  | IG | 87.80 | 9.40 | 152.00 | 12.53 | 73.12% |  |
| Self-perception<br>of Intelligibility | CG | 48.40 | 9.63 | 48.80 | 10.18 | 0.83% | .001 |
|  | IG | 42.00 | 8.90 | 72.40 | 7.19 | 72.38% |  |
| Word-level<br>intelligibility | CG | 0.62 | 0.10 | 0.62 | 0.10 | 0 % | .004 |
|  | IG | 0.59 | 0.10 | 0.85 | 0.06 | 43.92% |  |

**Legend:** Mean values (Mean) and percent change of average peak flow, self-perception of intelligibility and word-level intelligibility assessed before (Pre cRMT) and after (Post cRMT) the 28 day intervention. SE indicates standard error, p in between groups indicates significance of interaction between control group and intervention group. Degrees of freedom for average peak flow: $F(1,8)=24.36$ , for self-perception of intelligibility: $F(1,8)=22.89$ , and for word-level intelligibility: $F(1,8)=15.85$ .

Average peak expiratory flow increased by 4.66% in the control group, while the intervention group showed an increase of 73.12% (p=.001), corresponding to an improvement of 64.2 liters per minute. Self perception of intelligibility improved by 72.38% in the intervention group, compared to 0.83% (p=.001) in the control group, as assessed by VAS. Table 2 further shows that the speech intelligibility at word level improved by 43.92% in the intervention group, while it did not improve in the control group (0%)(p=.004). These results demonstrate significant improvements across outcomes in participants who performed cRMT compared to those who did not. While the measurements for the participants in the control group remained essentially the same over time, participants in the experimental condition (cRMT) showed significant improvement over time. Figure 1 illustrates the significant changes between groups post cRMT compared to before the study intervention. Figure 2 displays individual changes in each patient of the control and the intervention groups, respectively, underlining marked improvements in all intervention group patients across the three different assessments.

**Figure 1:**
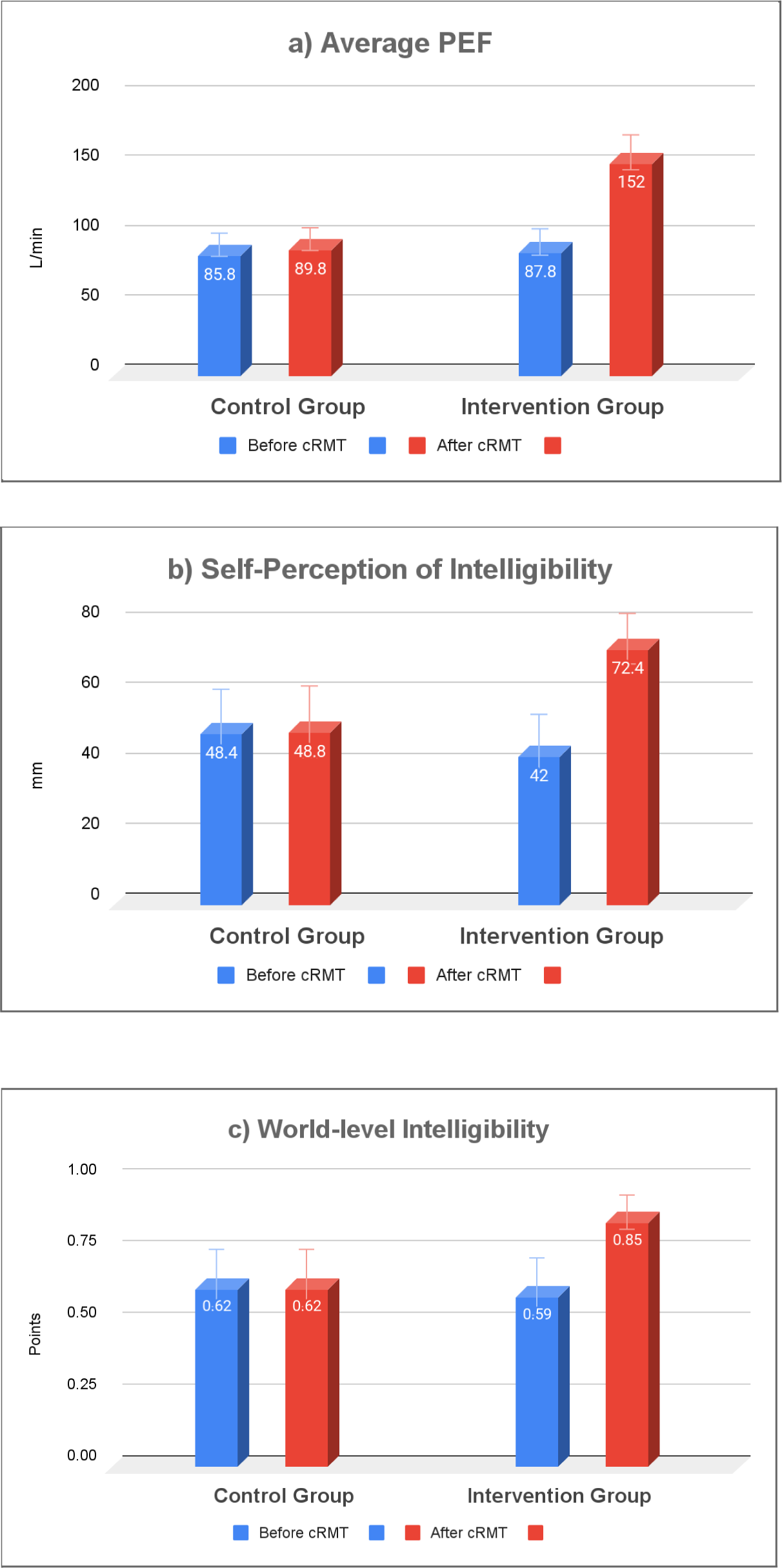
Illustration of cRMT-mediated group changes in peak expiratory flow (PEF) and intelligibility Four weeks of cRMT significantly improved average peak expiratory flow (PEF) (a), self-perception of intelligibility (b), word-level intelligibility (c) in the intervention group, but not in the control group. Error bars indicate standard error. Data is outlined in table 2.

**Figure 2:**
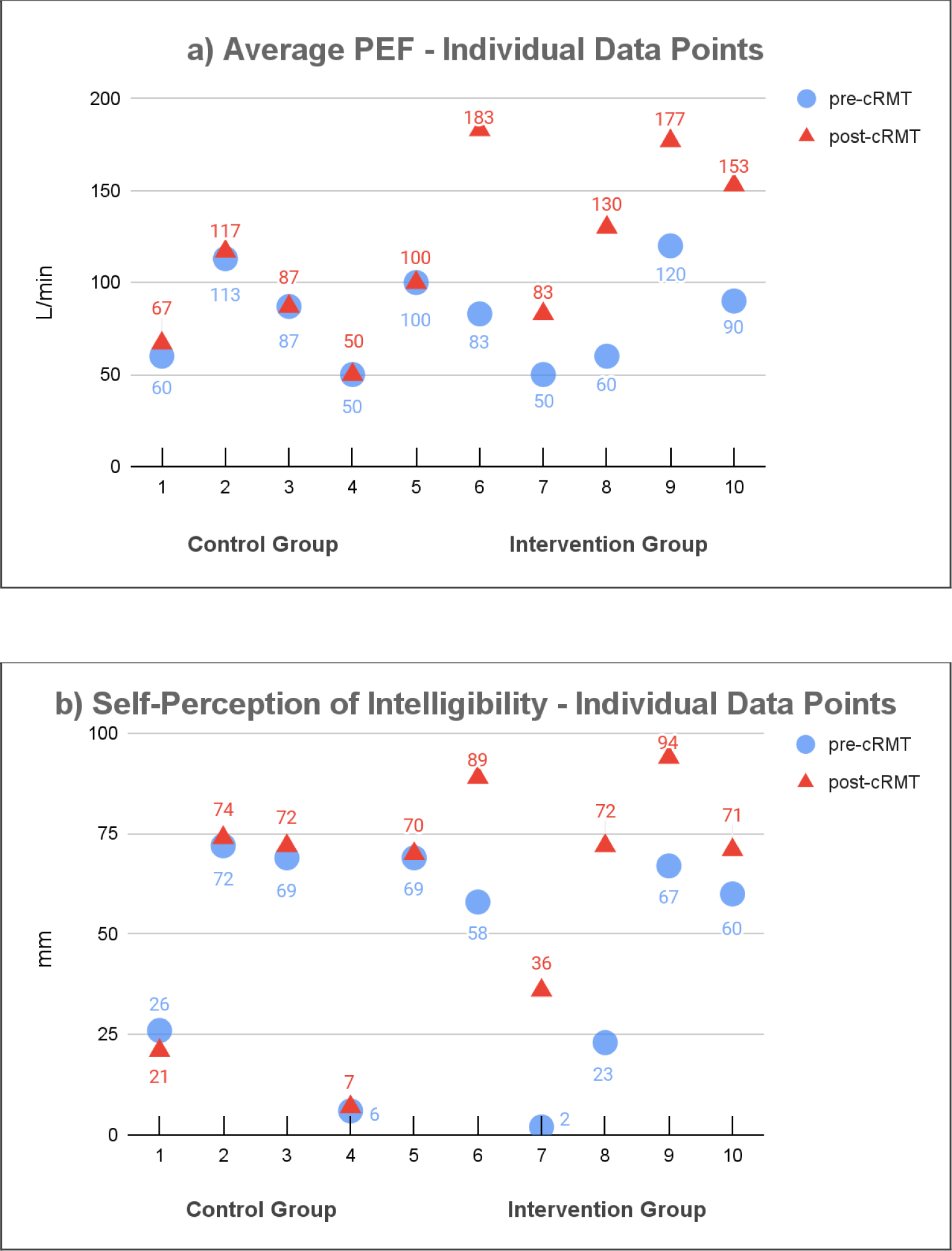
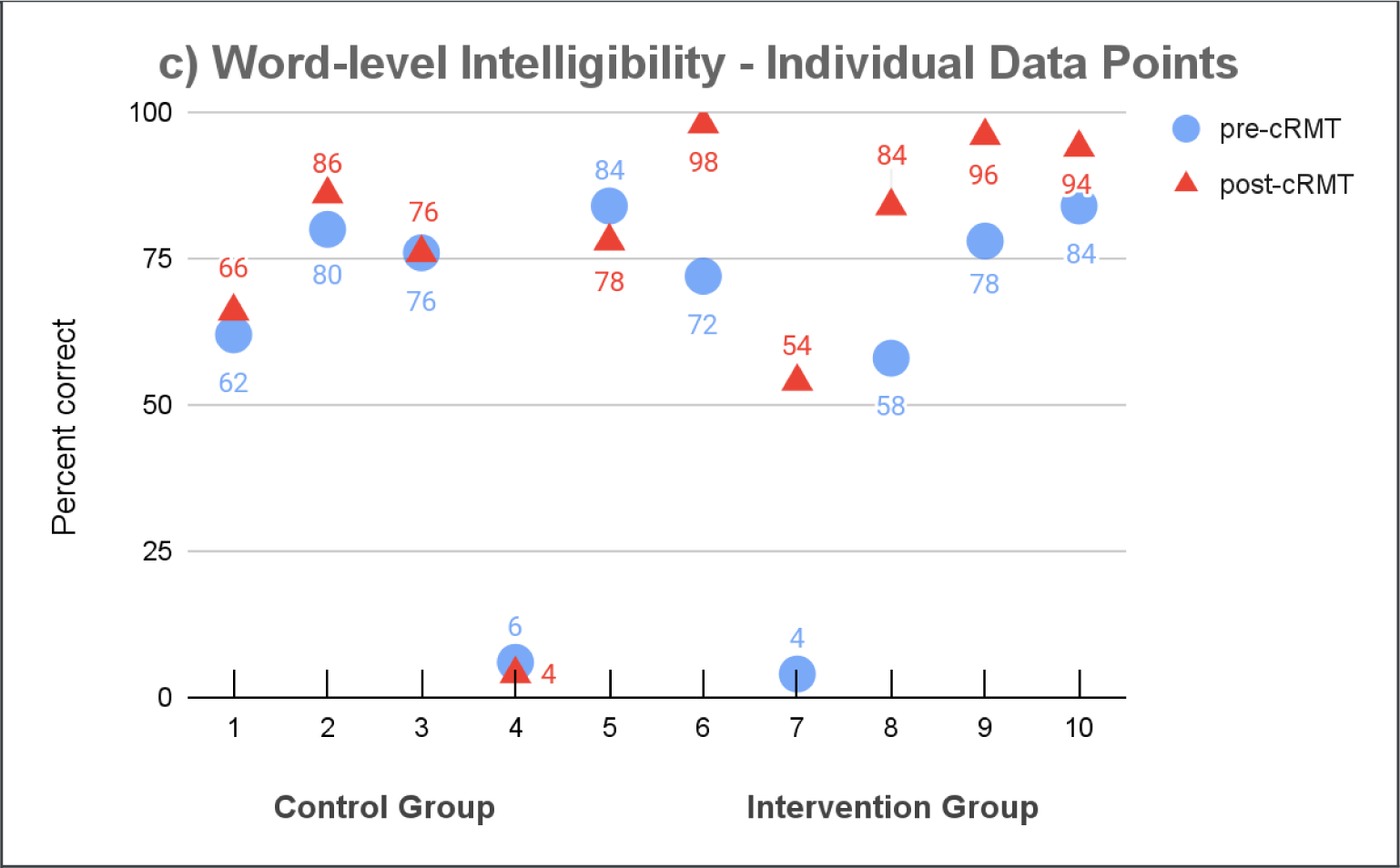
Illustration of cRMT-mediated changes on individuals Four weeks of cRMT improved peak expiratory flow (PEF)(a), self-perception of intelligibility (b), and word-level intelligibility (c) substantially in patients in the intervention group (6 to 10) compared to the control group (1 to 5).

## Discussion

This retrospective pilot study sought to determine the effectiveness of 4 weeks of 3 intensive brief daily sessions of cRMT on the speech intelligibility of stroke survivors with dysarthria. Compared to the control group whose data remained static, all of the participants in the experimental group showed positive gains after 4 weeks of cRMT in peak expiratory flow rate, self-perceived speech intelligibility, and speech intelligibility judged at the word level. As the sample size is limited, individual data points were illustrated (Figure 2), showing that the three parameters changes in every single patient in the intervention group, supporting the significance of the between group changes.

This study is unique in its attempt to determine the potential effects of cRMT on speech intelligibility. A recent study found some improvements in selected respiratory dynamics as well as measures of voice perturbation in stroke patients using both inspiratory and expiratory RMT, but no reports of speech intelligibility improvement were included (Liaw et al., 2020). The improvements in respiratory peak flow and intelligibility measures in this study suggest that cRMT can have a significant impact on patients with stroke-induced dysarthria.

The direct effect of cRMT on peak flow on the participants in the intervention group was not unexpected given the nature and intensity of the training and existing data from other studies (Arnold et al., 2021; Arnold & Bausek, 2020). It is likely that the experimental group participants experienced an increase in both inspiratory strength (resulting in higher initial lung volumes for the task), and expiratory strength (allowing for higher expiratory flow volume for the task). Both or either of the two effects could potentially explain the average 73% increase in peak flow for the experimental group.participants may have simply improved at the peak flow task. It is also possible that the changes were simply due to a practice effect after spending 4 weeks using the device. However, if it were merely a byproduct of practice effect, then the intervention group’s observed improvement should not have resulted in a higher increase than the control group. Furthermore, improvement in peak expiratory flow has also been observed in numerous other studies involving RMT across disorders ((Arnold et al., 2021; Arnold & Bausek, 2020; Bausek et al., 2019)). However, the accompanying improvements in both intelligibility measures suggest that the improvements in peak flow may indeed have been an experimental effect.

Both the experimental participants’ self-perceived intelligibility and their average word-level intelligibility as perceived by two blinded judges increased significantly after the 4-week cRMT training regimen. However, there was a larger (72%) increase in the perception of their own intelligibility than in their word-level intelligibility increase (44%) as perceived by the judges. This is likely due to the marked difference between intelligibility judged at the word level and at the sentence or conversational level. At the word level, no contextual cues are available to aid a listener in comprehension. It is assumed that the participants in the intervention group had an increase in word-level intelligibility because they were speaking with a higher initial lung volume after practising taking more forceful inhalations. It is difficult to predict from these data what their sentence or conversation-level intelligibility would have been; it may have increased similarly due to speaking with higher lung volumes and the addition of linguistic context, but those gains may have been limited due to decreasing lung volumes across longer utterances. It is interesting from a psychosocial standpoint that the experimental participants reported a 72% percent increase in the perception of their own intelligibility; as they felt that others understood them with less difficulty, this points to the validity of using cRMT for improving the functional intelligibility of dysarthric speech. Given the correlation between dysarthria and health related quality of life, it can be presumed that improvement in speech intelligibility has a positive impact on the overall quality of life and social interactions (Leite & Constantini, 2017; Spencer et al., 2020).

It is possible as well that the gains in both measures of intelligibility occurred due to an increase in the participants’ baseline subglottal pressure, and therefore vocal intensity, for speech. It is clear from the LSVT literature (Dromey et al., 1995; L. Ramig et al., 2018; L. O. Ramig & Dromey, 1996) that solely targeting loudness in treatment of hypokinetic dysarthria can have significant positive impact on not just the loudness of speech, but its rate and articulatory precision as well. While no speech tasks were directly targeted in this study, the carry-over effect from cRMT training to improved respiratory function and greater loudness during speech may be responsible for the positive gains seen in this study.

As impaired glottic closure is often a common finding following CVA, dysarthria, dysphonia, and dysphagia are frequent comorbidities (Martino et al., 2000, 2005; Park et al., 2020). Given the evidence of the effectiveness of cRMT on the treatment of dysphagia and dysphonia following single stroke (Arnold et al., 2021; Arnold & Bausek, 2020), the crossover treatment effects of cRMT may be cost-effective therapy when two or more of these physiological disorders exist. As other research has illustrated respiratory muscle weakness as a significant contributor to additional upper aerodigestive tract physiological disorders including dyspnea and dystussia (P. Nicholson & Covelli, 1989), for which RMT has emerged as an effective treatment option (Chaitow et al., 2014; McConnell, 2013), the crossover treatment effects of cRMT may also enhance cost-effectiveness when these or similar disorders co-occur.

Despite the small sample size and retrospective nature of this study, the magnitude of the observed gains in peak flow and intelligibility measures are rather striking. The fact that the intelligibility measures improved to such a degree in the experimental group suggests that cRMT could be a viable treatment option for making functional gains in communication for patients with dysarthria. The cRMT device used in this study is an accessible and inexpensive option for treating dysarthria since the task has a relatively low cognitive load. It is important to note that the nature of the training protocol used here, involving a high effort task employed within a high-intensity treatment regimen over a short period of time, adheres to muscle training principles of intensity, specificity and increasing load over time (McConnell, 2013). As the same principles of muscle strengthening in response to resistance training apply to the respiratory muscles, it is likely that increased respiratory muscle strength due to cRMT caused the speech intelligibility improvements observed in this study.

## Limitations

There are several possible limitations to this study that warrant caution in interpreting the results. The relatively small sample size and retrospective nature of the study are first on this list. The fact that the two groups were similar in their makeup of specific dysarthria types was beneficial for group comparisons; however, the fact that both experimental and control groups had a heterogeneous mix of dysarthria types makes it difficult to generalize the results of the study to specific populations of patients with dysarthria. The next potential limitation is that no assessment was made of the level of general debility or mobility post-CVA as it is possible that the intervention group had an overall physical advantage compared to the control group and may have more easily benefited from respiratory muscle training. Several limitations to the retrospective design of this study are derived from the limitations of the original data set. For example, it does not contain a SHAM control and the subjects can not be randomized between groups.

## Conclusion

This retrospective study analyzes the use of combined Respiratory Muscle Training (cRMT) with a hand-held device exercising both the muscles of inhalation and exhalation over a four week period revealing significant improvements in peak expiratory flow and speech intelligibility at the word level in the intervention group, as compared to the no treatment control group. In addition, this study also reveals superior patient perception with the intervention group in the domain of others understanding their speech. The present study therefore suggests the effectiveness and feasibility for the inclusion of cRMT in the treatment of persons who present with dysarthria after CVA. Our findings warrant a double blinded prospective, randomized sham-controlled study to verify these findings in a larger, more diverse population.

## Data Availability

All data can be made available upon request from SBA, LLC.

## Acknowledgements

The authors thank Sabine Elizabeth French, Ph.D., John F. Richardson, MS, CCC-SLP, Tom Slominski, MA, CCC-SLP, Sigfredo Aldarondo, M.D., FCCP, Tom Berlin, DHSc, RRT, Betsy Page, MA, CCC-SLP, Lawrence F. Johnson, M.D. for help with the statistical analysis, critical reading of the manuscript, and helpful comments.

## Disclosures and Contributions

RJA declares no conflict of interest. NB serves as independent Chief Scientist for PN Medical. CG declares no conflict of interest. TM declares no conflict of interest.

## Author’s contribution

Robert J. Arnold contributed to the study design, data acquisition, and interpretation of data, and writing of the manuscript.

Nina Bausek contributed to the interpretation of data, data analysis, and writing of the manuscript.

Christopher S. Gaskill contributed to the interpretation of data, and writing of the manuscript. Tarek Midani contributed to the writing of the manuscript.

All authors read and approved the final version of the manuscript.

## Notes

### Competing Interest Statement

CG declares no conflict of interest. NB serves as independent Chief Scientist for PN Medical. RJA declares no conflict of interest. TM declares no conflict of interest.

### Clinical Trial

This is not a prospective trial, therefore it was conducted under an IRB waiver granted by Western IRB.

### Funding Statement

No funding has been received for this study

### Author Declarations

Western IRB 1019 39th AVE SE, Suite 120 Puyallup WA 98374/2115 Phone: +1 360 252/2500 www.wirb.com

### Summary of Updates

This revision contains added figures for clarification of individual changes in the intervention and control groups.

